# Is mindfulness associated with lower pain reactivity and connectivity of the default mode network? A replication and extension study in healthy and episodic migraine participants

**DOI:** 10.1101/2022.01.18.22269473

**Authors:** Carly Hunt, Janelle E. Letzen, Samuel R Krimmel, Shana A.B. Burrowes, Jennifer A. Haythornthwaite, Patrick Finan, Maria Vetter, David A. Seminowicz

## Abstract

Formal training in mindfulness-based practices promotes reduced experimental and clinical pain, which may be driven by reduced emotional pain reactivity and undergirded by alterations in the default mode network (DMN), implicated in mind-wandering and self-referential processing. Recent results published in this journal suggest that mindfulness, defined here as the day-to-day tendency to maintain a non-reactive mental state in the absence of training, associates with lower pain reactivity, greater heat-pain thresholds, and resting-state DMN functional connectivity (FC) in healthy adults in a similar manner to trained mindfulness. The extent to which these findings extend to chronic pain samples and replicate in healthy samples is unknown. Using data from healthy adults (n = 36) and episodic migraine patients (n = 98) and replicating previously published methods, we observed no significant association between mindfulness and heat-pain threshold (*r* = -0.05, *p* = .80), pain intensity (*r* = -.02, *p* = .89) or unpleasantness (*r* = .02, *p* = .89), or pain catastrophizing (PC; *r* = .30, *p* = .08) in healthy controls, or between mindfulness and headache frequency (*r* = -.11, *p* = .26), severity (*r* = .03, *p* = .77), impact (*r* = -.17, *p* = .10) or PC (*r* = -0.09, *p* = .36) in patients. There was no association between DMN connectivity and mindfulness in either sample when probed via seed-based FC analyses. In post-hoc whole brain exploratory analyses, a meta-analytically derived DMN node (i.e., posterior cingulate cortex; PCC) showed connectivity with regions unassociated with pain processing as a function of mindfulness, such that healthy adults higher in mindfulness showed greater PCC-cerebellum FC. Collectively, these findings suggest that the relationship between mindfulness and DMN-FC may be nuanced or lacking in robustness, and cast doubt on mindfulness as a clinically meaningful protective factor in migraine.

**Perspective:** This study tested relationships between mindfulness and pain, pain reactivity and default mode connectivity in healthy adults and migraine patients. Findings cast doubt on mindfulness as an individual difference marker of the ability to cope with pain in healthy adults, and as a protective factor in episodic migraine.

## 1. Introduction

Emerging data suggest that mindfulness training reduces experimental ^47^ and clinical pain^39^, potentially via reduced cognitive-affective reactivity to pain vis-à-vis enhanced non-judgmental, present moment awareness ^25^. Alterations in the default mode network (DMN), a functionally connected network of brain regions underlying self-referential processing ^19^ and mind-wandering ^22^, have been implicated in the attentional changes that occur following mindfulness training ^4,9,11^. Further, multiple studies collectively suggest that trained mindfulness may favorably impact pain through a unique mechanism involving augmented attention towards sensory information and reduced emotional reactivity to pain.^12,14,26,48^ In comparison with mindfulness as a learned skill, less is known about mindfulness as a trait, or the tendency to maintain non-judgmental, present moment awareness in the absence of training ^1,3^. Suggestive of possible DMN involvement consistent with mechanisms underlying trained mindfulness, greater mindfulness in healthy controls associates with lower resting-state default mode connectivity ^15,33^, experimental pain ratings ^15,30,49^, and pain catastrophizing, a marker of emotional reactivity to pain ^15^ that associates with poorer functioning and reduced cortical grey matter volume in patients with migraine ^16,18^. The extent to which similar findings emerge in individuals living with chronic pain, particularly headache and migraine pain, remains unknown. Mindfulness inversely associates with pain severity in fibromyalgia ^10^ and in heterogeneous pain groups, including community-member undergraduates ^31^, adolescents ^34^, and adults presenting for chronic pain treatment ^29,38^. Furthermore, mindfulness consistently associates with pain-related variables such as pain interference, anxiety and depression, disability and health-related quality of life in chronic pain groups.^6,8,32,40^ However, null associations between pain severity and mindfulness have been reported in adolescents with pain ^42^, in knee osteoarthritis, ^23^ and in chronic low back pain.^6^ Whether mindfulness associates with reduced clinical pain in episodic migraine, a common and debilitating disorder ^24^, remains unclear. Furthermore, whether such potential pain reductions are undergirded by similar mechanisms as those underlying formally trained mindfulness in healthy controls remains unknown.

Given the above considerations, the present study investigated the degree to which the inverse associations between mindfulness and resting-state default mode connectivity, pain catastrophizing and heat-pain threshold in healthy controls (Harrison et al., 2019) extend to a chronic pain sample with episodic migraine. Secondly, we investigated the extent to which Harrison and colleagues’ (2019) findings replicate in a new sample of healthy controls. All participants were meditation naïve. We hypothesized that [1] higher mindfulness would associate with lower clinical pain (i.e., reduced headache frequency and headache pain reported via headache diary, and reduced self-reported headache impact), lower trait-like pain catastrophizing, and reduced DMN connectivity in patients and [2] reduced heat-pain threshold, experimental pain severity and unpleasantness, lower trait-like pain catastrophizing, and reduced DMN connectivity in controls.

## 2. Method

This was a secondary cross-sectional analysis of baseline data collected during a larger parent trial comparing an enhanced (i.e., 12-week) mindfulness-based stress reduction program (MBSR+) with an active control (i.e., Stress Management for Headache; SMH) condition ^39^. The full methodology of the parent project (R01AT007171) has been described elsewhere ^5,39^. Participants were recruited from 2014 to 2017, and all methods were approved by the Johns Hopkins School of Medicine and University of Maryland, Baltimore Institutional Review Boards. Here we detail methods pertinent to this secondary analysis. We used the Strengthening the Reporting of Observational Studies in Epidemiology (STROBE) checklist for this cross-sectional study, and include the checklist in Supplemental Material.

### Participants

Patient data were drawn from meditation naïve individuals (n = 98) who were randomized to either 12-week MBSR+ (n = 50) or HE (n = 48). Participants were included if they met criteria for the International Classification of Headache Disorders criteria for migraine with or without aura and had been living with migraine for at least 1 year. Participants were excluded if they reported severe psychiatric symptoms, opioid medication use, prior mindfulness experience, or engagement in any treatment anticipated to impact mindfulness including but not limited to cognitive-behavioral therapy, biofeedback, acupuncture, or massage therapy (see the <u>parent project Protocol</u> for full inclusion and exclusion criteria). Non-opioid medication use was permitted; however, patients were excluded if they were unwilling to stay on the same treatment regimen for at least 6 months, with the addition of rescue or abortive medications as needed. Healthy controls (n = 36) were matched to the first 36 enrolled patients based on age, sex, body mass index, and education. Healthy controls were eligible if they were free of acute and chronic pain conditions and lacking in migraine history. Demographic characteristics for both samples are shown in Table 1.

**Table 1.** Sample demographic characteristics.

|  |  |
| --- | --- |
| **Patients** |  |
| Total *n* | 98 |
| Age, mean (SD) | 38.2 (12.5) |
| Female, *n* (%) | 89 (91) |
| Headache Frequency (days), mean (SD) | 8.4 (2.9) |
| Headache Impact, mean (SD) | 60.5 (5.7) |
| Headache Pain Intensity (0-10), mean (SD) | 4.5 (1.6) |
| Mindfulness, mean (SD) | 140.53 (19.28) |
| Pain Catastrophizing | 11.9 (9.6) |
| Race, *n* (%) |  |
| Asian | 4 (4) |
| Black or African-American | 17 (17) |
| Multiracial | 3 (3) |
| Other | 2 (2) |
| White or European American | 71 (72) |
| Unknown or not reported | 1 (1) |
| College graduate, *n* (%) | 78 (80) |
| **Healthy controls** |  |
| Total *n* | 36 |
| Age, mean (SD) | 37.5 (12.9) |
| Female, *n* (%) | 32 (89) |
| Mindfulness, mean (SD) | 145.1 (19.0) |
| Pain Catastrophizing | 3.28 (5.79) |
| Race, *n* (%) |  |
| Asian | 2 (6) |
| Black or African-American | 8 (22) |
| White or European-American | 25 (69) |
| College graduate, *n* (%) | 32 (89) |
| Experimental Pain Severity (0-10), mean (SD) | 4.2 (2.3) |
| Experimental Pain Unpleasantness (0-10), mean (SD) | 4.3 (2.2) |
| Heat Pain Threshold (□), mean (SD) | 42.6 (3.1) |

### Measures

Mindfulness was measured with the Five Factor Mindfulness Questionnaire (FFMQ; Baer et al., 2006), which yields 5 subscale scores and a total score. Participants rate items on 5-point Likert scales (1 = *never or very rarely true*; 5 = *very often or always true*). Total scores were computed by taking the sum of each subscale score, and were used in analyses. Subscale scores were computed by taking the sum of subscale items. The 5 subscales include Non-Reactivity to Inner Experience (i.e., detaching from thoughts and emotions, allowing them to shift and change without becoming carried away by them), Observing (i.e., noticing internal and external experiences), Acting with Awareness (i.e., being attentive during activities, rather than acting on “autopilot”), Describing (i.e., expressing one’s experience using words), and Non-Judging of Inner Experience (i.e., relating to inner experience with an attitude of acceptance). Total scores demonstrated good internal consistency reliability in patients (Chronbach’s alpha = .85) and in controls (.87). Headache occurrence and severity were measured via 28-day electronic daily diary based on the National Institute of Neurological Disorders and Stroke preventive therapy headache diary. Headache frequency was prorated based on the number of completed diary days (i.e., to account for participants who completed fewer than 28 days), the proportion of headache days was computed (number of headache days/total number of diary days) and then multiplied by 28, yielding a continuous variable quantifying headache days. Headache severity was assessed with a 0-10 scale and was quantified as the average of all headache intensity ratings from the diary. Headache impact was measured with the Headache Impact Test (HIT-6), which assesses the effects of headaches on lifestyle ^20^; it has been validated in chronic and episodic migraine ^44^ and showed acceptable internal consistency reliability in this sample (Chronbach’s alpha =.71). Pain catastrophizing was measured with the pain catastrophizing scale ^41^, a 13-item measure which asks participants to rate how often they experience particular feelings and thoughts when experiencing pain (0 = *not at all*, 4 = *all the time*. Total scores demonstrated good internal consistency reliability in patients (Chronbach’s alpha = .91) and in controls (.93).

Experimental pain severity, unpleasantness and heat pain threshold in healthy controls were quantified through a quantitative sensory testing (QST) protocol using a 30 × 30 mm ATS probe (Medoc Pathway model, ATS, Medoc Advanced Medical Systems Ltd., Ramat Yishai, Israel. All stimuli were administered on the left volar surface of the forearm, and the probe was moved between stimulations. Heat-pain thresholds were quantified using the average of three heat pain threshold ratings, which were obtained by asking participants to signal via mouse-click the point at which they first felt pain sensation in response to rising temperatures, which began at a baseline temperature of 32°C and increased at a rate of 1.5°C/second. After heat-pain threshold testing, a set of 19 pseudorandom thermal stimuli were presented using a single ramp up and hold design. Participants verbally rated pain intensity and unpleasantness on 0 (no pain) to 10 (maximum pain imaginable) scales. Each simulation began with a baseline temperature of 32°C which then increased at a rate of 1.6°C per second until reaching the target temperature, and was then held constant for a duration of 6 seconds. There was a 6-second rest period between stimulations. The stimulus order was 40, 42, 44, 47, 41, 49, 41, 45, 48, 39, 49, 45, 48, 47, 45, 44, 43, 49, 47°C. The QST protocol has also been described in previous work by our group ^21^.

### Procedure

Patients completed 28 days of headache diary to determine eligibility for the parent project (4 – 14 headache days out of 28). Eligible patient participants, as well as healthy control participants, attended a magnetic resonance imaging (MRI) session that included quantitative sensory testing (QST) and completion of questionnaires.

### MRI Acquisition

Structural and functional MRI data were acquired at the University of Maryland, Baltimore Medical Imaging Facility with a Siemens 3T Tim Trio scanner. Either a 32-channel head coil (n = 22 healthy controls, 70 patients) or a Siemens 3T Prisma scanner with a 64 channel head coil (n = 14 healthy controls, 16 patients) were used; this change was made due to a scanner upgrade during data acquisition. We acquired a T1-weighted structural 3D MPRAGE scan, which was used in preprocessing steps (whole brain coverage, TR = 2300 ms, TE=2.98 ms, voxels = 1.00 mm isotropic). We also acquired an eyes-open, resting-state scan using EPI while participants fixated on a plus sign (whole brain coverage, TR = 2000 ms, TE = 28 ms, voxels 3.4 × 3.4 × 4.0 mm, slices = 40, duration = 300 TR).

### MRI Analysis Overview

The present study aimed to determine the reproducibility of results by Harrison and colleagues (2019) in a new sample of healthy controls and extend these results to a sample of adults with episodic migraine. We further conducted post-hoc exploratory analyses to examine the reproducibility of results by Parkinson and colleagues (2019) in healthy controls and their extension in adults with episodic migraine. For the exploratory analyses, we only aimed to examine the reproducibility of Parkinson et al.’s findings in the context of the DMN, which is consistent with our study aim. In the below sections, selection of the study’s regions of interest (ROIs) and masks, preprocessing, denoising, and first-level analyses were applied across the entire sample. Group-level analyses, however, were conducted separately for the sample of healthy controls and the sample of adults with episodic migraine. Of the 98 participants randomized, resting-state fMRI data from 93 participants adequately passed quality control measures and were included in the present study.

### Regions of Interest and Mask Selection

#### Reproduction of Harrison et al. (2019)

In the previous work by Harrison and colleagues, functional connectivity of the DMN was measured using seed-based functional connectivity analyses. Two regions of interest (ROIs) were selected based on those in their report. In the present study, we used the Wake Forest University Pickatlas toolbox ^27,28^ to construct lateralized precuneus seeds by projecting 2-mm spheres around the following coordinates in Montreal Neurological Institute space: X=-8 or 8, Y=-64, Z=18. These ROIs were used as the main seeds in seed-based functional connectivity analyses. Further, Harrison and colleagues applied a meta-analysis mask to second-level seed-based functional connectivity analyses to examine DMN functional connectivity with pain-related brain regions as it associated with mindfulness. To recreate this meta-analysis mask, we followed steps by Harrison and colleagues that included conducting a term-based meta-analysis from the Neurosynth database ^45^ for the keyword “pain” and downloading the resultant association test, binarized mask.

#### Additional Analyses

Based on the results of the main reproduction analyses, we conducted two sets of additional analyses to further test the reproducibility of mindfulness as a predictor of DMN FC with pain-related regions. These analyses required separate sets of ROIs than those noted above.

First, we conducted exploratory ROI-to-ROI FC analyses in our dataset between Harrison et al.’s (2019) precuneus seeds and areas that were identified as significantly associated with the precuneus seeds as a function of mindfulness by Harrison et al. ROIs were created using the Pickatlas toolbox by drawing 6mm spheres around the following peak coordinates reported in Harrison et al.: [1] right parietal/motor/somatosensory cortex (32, -18, 38), [2] left parietal/motor/somatosensory cortex (-10, -46, 52), [3] left parietal/somatosensory cortex (14, - 32, 44), [4] medial prefrontal cortex/perigenual ACC (-2, 44, 6), [5] left superior frontal gyrus/premotor (-10, 36, 54), [6] right superior frontal gyrus/premotor (6, 22, 66), and [7] posterior cingulate cortex/precuneus (-10, -50, 26).

Second, we conducted exploratory seed-to-voxel FC analyses using two key DMN nodes [PCC and ventromedial prefrontal cortex (vmPFC)] derived from the Neurosynth database. To identify these nodes, “default mode” was used to conduct a term-based meta-analysis that included 777 studies with 26,256 activations. Using the Pickatlas toolbox, 6mm spheres were drawn around activation peaks at the following coordinates: PCC (0, -52, 26; z-score for “default mode”=15.92) and vmPFC (0, 50, 4, z-score for “default mode”=7.61).

### MRI Data Processing and Denoising

Preprocessing was completed in SPM12 and included slice timing correction, realignment (motion correction), coregistration of the T1 to the mean functional image, segmentation of the T1, normalization of functional images with interpolation to 2 × 2 × 2-mm voxels, and smoothing with a 6 mm full width at half maximum (FWHM) Gaussian kernel. Data were visually inspected at each preprocessing stage for quality control. Motion regression was based on framewise displacement using custom scripts so that participants with FrameWise Displacement Arithmetic Mean greater than 0.3 were removed 13T(Power et al., 2012, 2014, 2015)13T.

Preprocessed resting-state data were then entered in CONN toolbox (version 17f; 36TUhttp://www.nitrc.org/projects/connU36T) for additional processing. Global signal was not removed from our analyses based on ongoing controversy (Murphy and Fox, 2017). The aCompCor algorithm (Behzadi et al., 2007; Muschelli et al., 2014) was used to control for white matter (WM) and cerebral spinal fluid (CSF) confounds. Eroded WM and CSF masks that did not include external or extreme capsules were used since we did not remove global signal (Power et al., 2017). Denoising further included removal of realignment parameters along with first-order derivatives of these parameters, simultaneous bandpass filtering between 0.008 and 0.09 Hz, linear detrending, and despiking after these regression steps (Patel et al., 2014). Denoised data were visually inspected for quality control to ensure that processing resulted in a normalized distribution of connectivity values for each participant.

### First-Level Analyses

#### Reproduction of Harrison et al. (2019)

Following denoising, participant-level data underwent first-level analysis in CONN. Averaged timeseries data from all voxels within each precuneus seed ROIs were extracted. Extracted values were entered as a regressor of interest in whole-brain FC analyses.

#### Additional Analyses

For the ROI-to-ROI FC exploratory analyses, individual-level averaged timeseries from all voxels inside of each 6mm ROI sphere were respectively extracted and entered as regressors of interest. For the seed-to-voxel FC exploratory analyses, individual-level averaged timeseries data from all voxels inside of each 6mm Neurosynth-based ROI sphere were respectively extracted and entered as regressors of interest in whole-brain FC analyses.

### Self-reports

Means and standard deviations were calculated for demographic variables, primary measures and covariates. Pearson’s correlations were used to test the associations between headache frequency, headache impact, and trait-like pain catastrophizing in patients, and between mindfulness, pain catastrophizing, and experimental pain (severity, unpleasantness and sensitivity) in healthy controls. Consistent with Harrison and colleagues (2019), we also entered pain catastrophizing and heat pain threshold into a multiple linear regression model predicting mindfulness, in order to test their unique associations with mindfulness. Missing data were handled using listwise deletion in these models.

### Group-Level Functional Connectivity Analyses

#### Reproduction of Harrison et al. (2019)

First-level contrast maps were entered into separate group-level analyses for the episodic migraine and healthy control samples. We initially examined the extent to which the precuneus seeds resulted in DMN FC in our sample by entering each seed into two separate, group-level models as regressors of interest. Then, participants’ FFMQ 5-factor total scores were added into each of these group-level model as a regressor of interest, so that significant positive clusters represented areas of greater FC with the precuneus seed as a function of greater mindfulness.

#### Additional Analyses

As previously noted, we conducted three follow-up sets of analyses based on the findings from the reproduction analyses described above: [1] ROI-to-ROI FC exploratory analyses using ROIs based on the seeds and identified clusters from Harrison et al. and FFMQ scores as a regressor of interest and [2] seed-to-voxel FC exploratory analyses using Neurosynth-based DMN nodes and FFMQ scores as a regressor of interest.

First, the ROI-to-ROI FC exploratory analyses were conducted to determine the extent to which significant associations that were identified by Harrison et al. were observable in our samples of healthy controls and individuals with episodic migraine. Individual-level Pearson’s r- to-z associations between each precuneus seed ROI and the seven ROIs based on Harrison et al.’s findings were entered in group-level models with FFMQ scores as a regressor of interest. Additionally, the same model was repeated substituting PCS total scores in place of FFMQ scores to determine whether the association between pain catastrophizing and mindfulness-associated DMN FC was observable in our samples of healthy controls or patients with episodic migraine.

Second, the Neurosynth-based seed-to-voxel FC exploratory analyses were conducted to determine the extent to which using robust DMN seeds elicited similar FC patterns – as a function of FFMQ scores – to those reported in Harrison et al. Averaged timeseries across all of the voxels within the PCC and vmPFC ROIs, respectively, were extracted and entered into group-level models as regressors of interest, both with and without FFMQ scores as an additional regressor of interest. Consistent with analyses in Harrison et al., averaged timeseries across voxels in significant clusters that were identified at the group-level were extracted for each individual, so that Pearson’s correlations could be conducted between these FC values and PCS total scores.

#### Thresholding

Thresholds were consistent across all FC analyses and used the standard settings for two-tailed, parametric cluster-based inferences (i.e., Gaussian Random Field theory)^43^ in the CONN toolbox: voxel threshold = *p<*.001 uncorrected and cluster threshold = *p<*.05 cluster-size *p*FDR corrected.

## 3. Results

### Descriptive Data

Table 1 summarizes demographic characteristics, mindfulness, pain catastrophizing and pain variables for patients and healthy controls. All participants had complete data on pain, pain catastrophizing and mindfulness data, except one migraine patient who was missing on mindfulness.

### Self-Reported Patient Data: Clinical Pain and Pain Catastrophizing

Headache frequency, severity and impact distributions approximated normality and contained no outliers (+/- 3 *SD*). One outlier in pain catastrophizing was detected using the +/- 3 *SD* criterion and was removed. Mindfulness was not associated with headache frequency, headache pain severity, headache impact, or pain catastrophizing (*p*’s > .05) as shown in Table 2.

**Table 2.** Correlations between mindfulness, headache characteristics and catastrophizing in patients (*n* = 98)

| Variable | 1 | 2 | 3 | 4 |
| --- | --- | --- | --- | --- |
| 1. Mindfulness |  |  |  |  |
| 2. Headache Frequency | -.11 |  |  |  |
| 3. Headache Severity | .03 | -.08 |  |  |
| 4. Headache Impact | -.17 | .003 | .39** |  |
| 5. Pain Catastrophizing | -.09 | .11 | .19 | .41** |

*Note.* \* indicates $p < .05$ . \*\* indicates $p < .01$ .

### Self-Reported Healthy Control Data: Experimental Pain and Pain Catastrophizing

All experimental pain variable distributions approximated normality and contained no outliers (+/- 3 *SD*). One outlier in pain catastrophizing was detected using the +/- 3 *SD* criterion and was removed. Mindfulness was not associated with heat pain threshold, experimental pain intensity, experimental pain unpleasantness, and pain catastrophizing (*p*’s > .05) as shown in Table 3. When entered into a multiple linear regression model, neither pain catastrophizing (*b* = -.13, *t*(33) = -.21, *p* = .83) nor heat pain threshold (*b* = -.36, *t*(33) = -.32, *p* = .75) was associated with mindfulness (see Table 4).

**Table 3.** Correlations between mindfulness, pain and pain catastrophizing in healthy controls (*n* = 36)

| Variable | 1 | 2 | 3 | 4 |
| --- | --- | --- | --- | --- |
| 1. Mindfulness |  |  |  |  |
| 2. Heat-pain threshold | -.04 |  |  |  |
| 3. Pain Severity | -.02 | -.55** |  |  |
| 4. Pain Unpleasantness | .02 | -.50** | .93** |  |
| 5. Pain Catastrophizing | .30 | -.37* | .08 | .09 |

*Note.* $M$ and $SD$ are used to represent mean and standard deviation, respectively. \* indicates $p < .05$ . \*\* indicates $p < .01$ .

**Table 4.** Joint effects of heat-pain threshold and pain catastrophizing on mindfulness

| *Predictors* | Mindfulness |  |  |
| --- | --- | --- | --- |
|  | *Std. Estimates* | *Std. CI* | *p* |
| Intercept | 0.00 | -0.35-.35 | 0.003 |
| Heat-pain threshold | -0.06 | -0.44-0.32 | 0.75 |
| Pain catastrophizing | -0.04 | -0.42-34 | 0.83 |

*Notes.* Model $R^2$ : 0.00

### Primary Imaging Analyses: Reproduction of Harrison et al. (2019)

Figure 1 along with Tables S1 and S2 demonstrate seed-based functional connectivity analyses between the left and right precuneus seeds for healthy controls and adults with episodic migraine without examining FFMQ scores as a regressor of interest. Unlike findings from Harrison et al., the precuneus seed did not generate a hallmark pattern of DMN FC in our sample. When FFMQ scores were entered in the model, no clusters emerged as significantly associated with either precuneus seed in either participant set. Further, restricting the search space within the Neurosynth “pain” meta-analysis mask, as conducted in Harrison et al., did not yield significant clusters.

**Figure 1.**
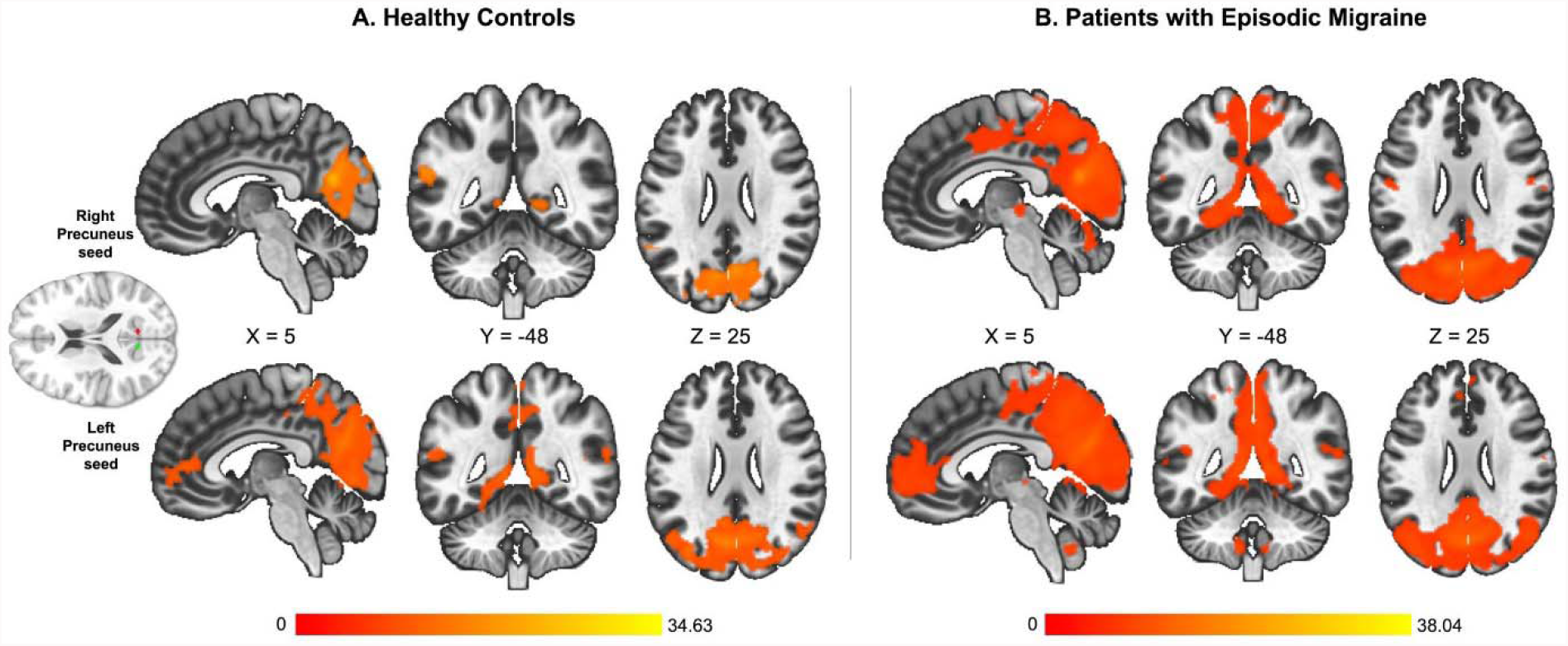
Seed-based functional connectivity analyses between the left and right precuneus seeds for healthy controls and adults with episodic migraine without examining FFMQ scores as a regressor of interest. Color bars represent t-statistics for connectivity maps.

### Secondary Imaging Analyses: ROI-to-ROI Exploratory Analyses

Without accounting for FFMQ scores, significant ROI-to-ROI associations in both patient and control samples were observed between the Harrison et al., precuneus seeds and the seven ROIs based on clusters identified in their main analysis (Table 5). In both the healthy control and patient samples, however, these effects no longer reached statistical significance after entering FFMQ total scores or PCS total scores in the model.

**Table 5.**
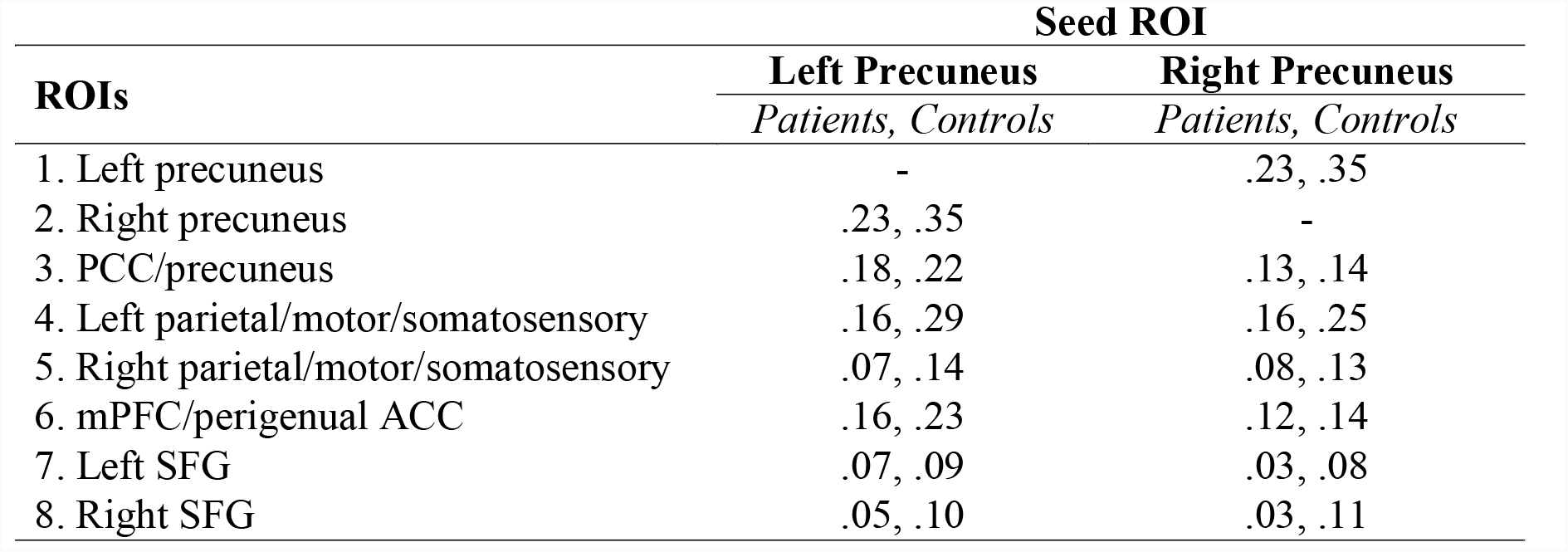
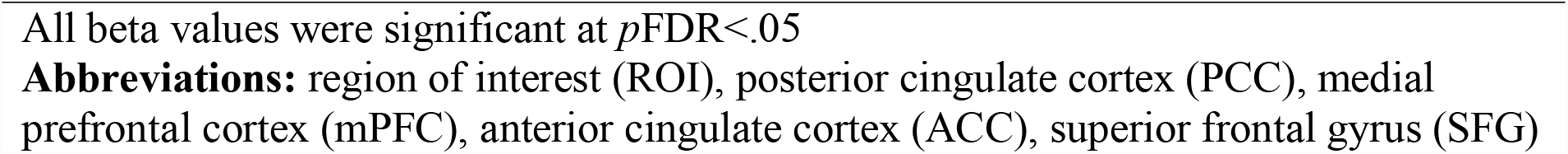
Beta values for ROI-to-ROI functional connectivity analyses among precuneus seeds derived from Harrison et al., 2019 and ROIs created around clusters identified in primary analyses by Harrison et al., 2019 without accounting for FFMQ or PCS total scores

| ROIs | Seed ROI |  |
| --- | --- | --- |
|  | Left Precuneus | Right Precuneus |
|  | *Patients, Controls* | *Patients, Controls* |
| 1. Left precuneus | - | .23, .35 |
| 2. Right precuneus | .23, .35 | - |
| 3. PCC/precuneus | .18, .22 | .13, .14 |
| 4. Left parietal/motor/somatosensory | .16, .29 | .16, .25 |
| 5. Right parietal/motor/somatosensory | .07, .14 | .08, .13 |
| 6. mPFC/perigenual ACC | .16, .23 | .12, .14 |
| 7. Left SFG | .07, .09 | .03, .08 |
| 8. Right SFG | .05, .10 | .03, .11 |

All beta values were significant at $p\text{FDR} < .05$
**Abbreviations:** region of interest (ROI), posterior cingulate cortex (PCC), medial prefrontal cortex (mPFC), anterior cingulate cortex (ACC), superior frontal gyrus (SFG)

### Secondary Imaging Analyses: Neurosynth-Based DMN Seed-to-Voxel Exploratory Analyses

Tables S3 and S4 along with Figure 2 detail the voxel clusters with significant functional connectivity to NeuroSynth-derived PCC and vmPFC seeds in healthy controls and patients without accounting for FFMQ total scores. When FFMQ scores were added as a regressor of interest, the Neurosynth-derived PCC node of the DMN was associated with one cluster at the whole-brain level in the healthy control sample only. Specifically, the PCC node was positively associated with a left lateralized cluster in the cerebellum (Table 6, Figure 3). PCS total scores were not associated with PCC-cerebellum FC values (r=.29, *p*=.09) which might be attributable to the floor effect of PCS total scores among healthy controls in this study.

**Figure 2.**
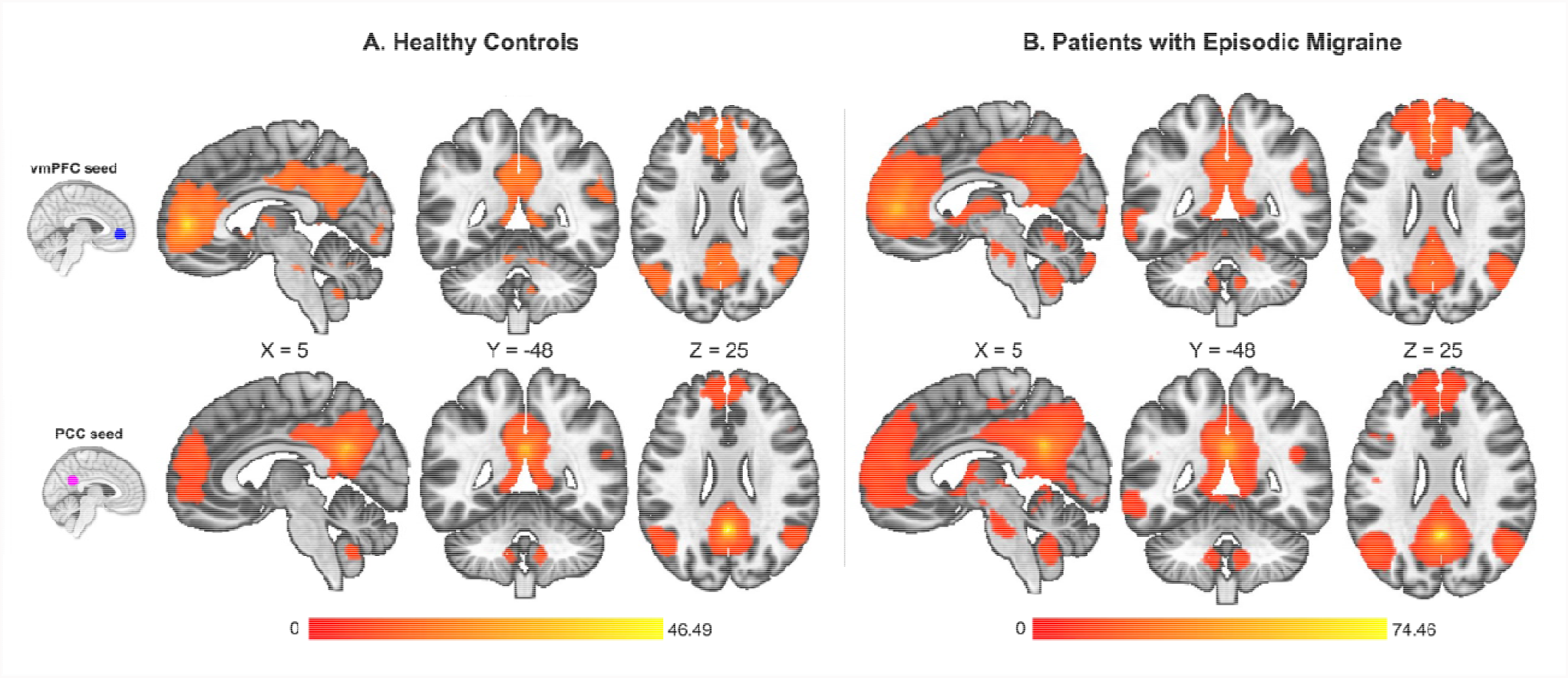
Voxel clusters with significant functional connectivity to NeuroSynth-derived PCC and vmPFC seeds in healthy controls and patients without accounting for FFMQ total scores. Color bars represent t-statistics for connectivity maps.

**Table 6.** Regions showing significant functional connectivity with the NeuroSynth-derived PCC seed as a function of mindfulness (FFMQ total scores) in healthy controls

| Seed | Cluster Coordinates | Cluster Size | Cluster Size $p$FDR | Cluster Regions |
| --- | --- | --- | --- | --- |
| PCC | -48 -72 -38 | 135 | .04 | Left cerebellum crus I/II |

**Figure 3.**
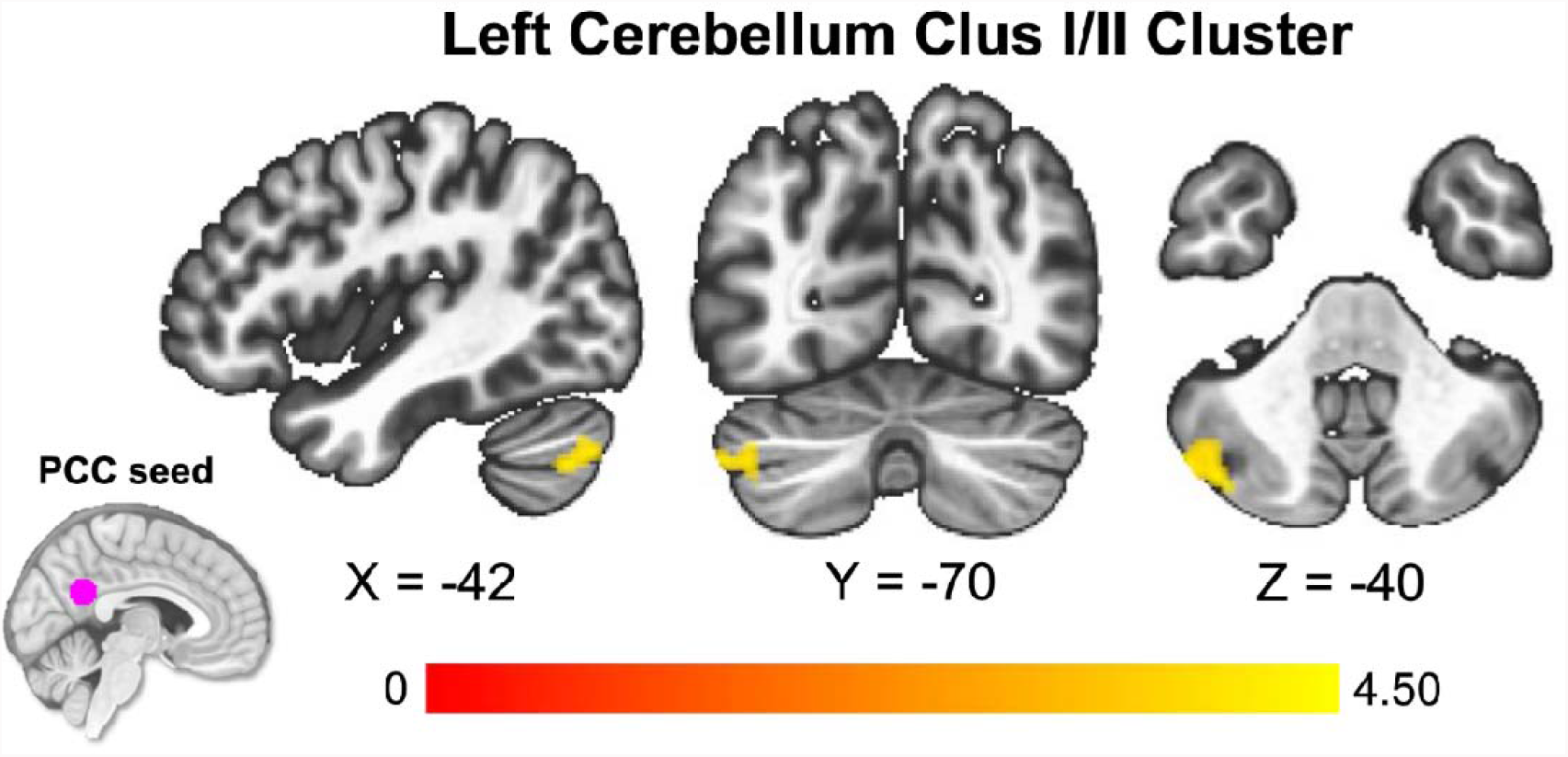
Significant associations emerging from Neurosynth-based DMN seed-to-voxel exploratory analyses in healthy controls. Color bars represent t-statistics for connectivity maps.

In the patient sample, Neurosynth-derived PCC and vmPFC nodes of the DMN were not significantly associated with any clusters at the whole-brain level as a function of FFMQ sum scores.

### Sensitivity analyses

Some psychometric work suggests that a 4-factor model, without the Observing FFMQ facet, provides the best fit for meditation naïve respondents ^2^. We thus tested associations between 4-factor FFMQ scores and connectivity in the aforementioned networks, as well as with clinical pain outcomes. In these analyses, mindfulness was not associated with headache frequency, severity, impact or pain catastrophizing in patients, and was unassociated with experimental pain and pain catastrophizing in healthy controls (*p*’s > .05). Substitution of the 4-factor sum score in place of the 5-factor sum score in the neuroimaging models did not change primary analysis results.

### Post-hoc exploratory analyses

#### Subscale Analyses in Patients

In a post-hoc analysis, we conducted subscale analyses by correlating FFMQ subscale scores with PCS subscale scores, clinical pain variables, and DMN connectivity (full results shown in Supplemental Material, Table S5). Notably, a significant, inverse correlation emerged between the non-judging facet of mindfulness and the magnification subscale of pain catastrophizing (*r* = -.21, *p* < .05). Also, there was a significant, inverse association between the non-judging facet of mindfulness and headache frequency (*r* = -.29, *p* < .01). For the non-judging subscale, greater scores were associated with greater vmPFC FC to DMN regions (e.g., angular gyrus, precuneus/PCC) and greater PCC FC to a cluster spanning anterior cingulate and paracingulate gyri (full results available in Supplemental Material, Table S6). No other subscales associated with DMN FC data in patients.

#### Experimental Pain in Patients

We explored the associations between mindfulness and experimental pain variables in patients, as the patient sample underwent the same experimental pain testing protocol as the healthy control sample. In patients, mindfulness was unassociated with heat pain threshold (*r* = .01, *p* = .98), experimental pain intensity (*r* = .15, *p* = .17) and experimental pain unpleasantness (*r* = .09, *p* = .39).

#### Subscale Analyses in Healthy Controls

We correlated FFMQ subscale scores with PCS subscale scores and experimental pain severity, unpleasantness, and heat pain thresholds. No significant correlations emerged between FFMQ subscale scores and experimental pain variables, or between FFMQ subscale scores and PCS subscale scores (*p*’s > .05); full results shown in Supplemental Material, Table S7. FFMQ subscale scores did not show associations with DMN connectivity; full results shown in Supplemental Material, Table S8.

## Discussion

The principal finding of this study is that the inverse association between mindfulness and DMN connectivity observed in a healthy sample of 40 healthy participants (Harrison et al., 2019) did not replicate in our sample of 36 heathy adults, or extend to a much larger (n = 93) chronic pain sample. Specifically, in contrast to hypotheses, we did not observe any significant association between DMN connectivity and mindfulness in healthy controls or in patients when probed via seed-based functional connectivity analyses. In addition, pain catastrophizing did not associate with significant patterns of seed-based functional connectivity. In a consistent manner, mindfulness was not associated with experimental pain in healthy controls, clinical pain in patients, or pain catastrophizing in either group. These findings challenge the notion that mindfulness might serve as an individual difference marker of the ability to cope with pain in healthy subjects, and that there is a shared neurobiological mechanism between trait and state mindfulness. Further, our null findings in patients challenge the robustness of mindfulness as a protective factor in the context of chronic pain, and episodic migraine in particular.

In post-hoc exploratory analyses, we did observe that the meta-analytically derived PCC node of the DMN was associated with one cluster at the whole brain level as a function of mindfulness in healthy controls. Specifically, heathy individuals higher in mindfulness evidenced greater connectivity between the PCC and a left lateralized cluster in the cerebellum. Pain catastrophizing was not associated with these patterns of connectivity. These findings suggest that aspects of DMN FC could be related to mindfulness in healthy subjects, and that there may be variability in which DMN nodes demonstrate this association. However, given that (1) we did not observe FC patterns involving regions relevant to pain processing in relation to mindfulness, (2) pain catastrophizing did not associate with DMN FC, and (3) mindfulness was unassociated with experimental pain, these findings additionally fail to support the robustness of mindfulness as a meaningful individual difference marker of pain processing or reactivity in healthy adults.

Additional post-hoc exploratory analyses included subscale analyses, which must be interpreted with caution due to multiple comparisons. For the most part, subscale analyses also failed to provide evidence of associations between DMN connectivity, pain and pain reactivity. However, higher levels of non-judging of inner experience were associated with reduced headache frequency, lower tendency to magnify the experience of pain, and greater functional connectivity to regions of the DMN using the meta-analytically derived vmPFC seed and to anterior cingulate/paracingulate gyri using the meta-analytically derived PCC seed in patients. Tentatively, it is possible the non-judging facet of mindfulness is a somewhat meaningful protective factor in the context of migraine, but replication of this finding in other samples is necessary.

Relatively little prior work is available to contextualize the present findings. One recent study in migraine reported that mindfulness buffered negative affective reactivity to pain on a day-to-day basis, but did not report on direct relations between mindfulness and pain ^7^. Mindfulness has been inversely associated with pain intensity in heterogeneous pain groups, including undergraduate students ^31^, adolescents ^34^ and adults presenting for chronic pain treatment ^29^; although, consistent with our results, null associations between mindfulness and pain severity have been reported ^42^. Although training in mindfulness-based stress reduction has been shown to reduce headache frequency and impact in migraine ^39^, mindfulness as a dispositional characteristic appears less meaningful in this context. Perhaps the active and volitional use of mindfulness strategies as acquired through systematic training is of greater clinical importance and neurobiologically significant ^47,48 4,13,17,46^ than the presence of a mindfulness-like trait occurring in the absence of training.

Although prior findings in a similarly sized healthy sample led us to hypothesize that DMN connectivity would be associated with mindfulness, some methodological details, including differences in scanning parameters, study contexts, toolboxes and processing pipelines distinguish the present study from prior work (Harrison et al., 2019). However, we did replicate the individual and group-level analyses used in Harrison et al (2019), and we were able to robustly identify the DMN despite these differences in methods. The relationship between mindfulness and DMN connectivity may be nuanced, perhaps emerging only in certain individuals or circumstances. Less than half of the European sample in Harrison and colleagues (2019) was female, whereas the majority (86%) of healthy control participants in the present study were female, and living in the United States. Further research might explore whether gender plays a moderating role on the associations between DMN connectivity, pain catastrophizing and mindfulness. Other demographic characteristics like age, racialized identity, and education level were not reported in Harrison et al (2019), making it difficult to understand additional individual difference factors. More broadly, it is likely that there were unmeasured characteristics that differed systematically between the two samples, contributing to non-replication. As with all new areas of research, a certain amount of non-replication is expectable, and our findings encourage further investigations of these questions in new samples, including other pain conditions.

In terms of limitations, it should be noted that the current patient sample was waiting to start a stress reduction program that included meditation for some, and therefore factors like treatment expectations and hypervigilance could have biased how patients reported their mindfulness, pain or pain catastrophizing. Moreover, our healthy control sample was relatively small. Future research should continue to characterize the importance of mindfulness as a protective factor in the context of chronic pain. Replication of these findings in additional healthy and chronic pain groups, and investigation in more gender and ethnically diverse samples, would be informative. For example, it could be that there are currently unidentified subgroups of patients who may reap a protective benefit from untrained mindfulness. Another intriguing question is whether mindfulness measures tap the same psychological construct prior to and following formal exposure to mindfulness-based concepts, as formal training might change how individuals understand and reflect on the items. This issue might be explored in future longitudinal studies.

Of note, while the methods used in Harrison and colleagues (2019) and ours were quite similar, there were some differences in the quantitative sensory testing procedures. The major differences were as follows: 1) the stimulus ramp rate (.5 degrees/second in Harrison et al. vs 1.5 degrees/second in ours); 2) Harrison et al. (2019) also incorporated the method of levels, which could have led to more reliable measurements of heat pain threshold; 3) Harrison et al. (2019) stimulated the lower right calf, while we stimulated the left volar forearm; 4) Harrison et al (2019) used a visual analogue scale, whereas we used a verbal numerical rating scale; 5) Harrison et al. (2019) reported heat-pain threshold data only, whereas we included ratings of pain intensity and unpleasantness in response to a series of pseudorandom stimuli, in addition to heat-pain threshold.

## Conclusions

In conclusion, the present analyses did not identify meaningful associations between mindfulness and pain, pain catastrophizing or default mode connectivity in healthy adults or episodic migraine patients, which contrasts with prior findings ^15^. Our results suggest that untrained mindfulness may not be an informative individual difference marker of the ability to cope with pain in healthy adults, or as a protective factor in episodic migraine.

## Supporting information

Supplemental Material (including STROBE checklist)

## Data Availability

All data produced in the present study are available upon reasonable request to the authors.

## Acknowledgments

This work was supported by National Institutes of Health awards T32NS070201 (CAH) and R01AT007176 (DAS).

