## Supplemental Material (including STROBE checklist) for "Is mindfulness associated with lower pain reactivity and connectivity of the default mode network? A replication and extension study in healthy and episodic migraine participants"

*Table S1.* Voxel clusters with significant functional connectivity to right and left precuneus seeds (based on parameters from Harrison et al., 2019) in healthy controls without accounting for FFMQ total scores

| **Precuneus Seed** | **Cluster Coordinates** | **Cluster Size** | **Cluster Size *p*FDR** | **Cluster Regions** |
| --- | --- | --- | --- | --- |
| Right | 8 -64 18 | 5172 | <.001 | Right precuneus/PCC  Bilateral cuneal cortex  Bilateral intracalcarine cortex  Bilateral supracalcarine cortex  Bilateral lingual gyrus  Bilateral lateral occipital cortex, superior division  Left lateral occipital cortex, inferior division  Left supramarginal gyrus, posterior division  Left angular gyrus  Left middle temporal gyrus, temporooccipital  Right occipital pole  Vermis VI/IV |
| Left | -8 -64 18 | 13331 | <.001 | Left precuneus/PCC  Bilateral cuneal cortex  Bilateral intracalcarine cortex  Bilateral supracalcarine cortex  Bilateral lingual gyrus  Bilateral lateral occipital cortex, superior division  Bilateral lateral occipital cortex, inferior division  Bilateral angular gyrus  Bilateral supramarginal gyrus, posterior division  Bilateral occipital fusiform gyrus  Bilateral occipital pole  Bilateral temporal occipital fusiform cortex  Bilateral middle temporal gyrus, temporooccipital  Bilateral precentral gyrus  Bilateral postcentral gyrus  Left temporal fusiform cortex, posterior division  Left parahippocampal gyrus, posterior division  Left hippocampus  Left planum temporale  Bilateral cerebellum/vermis IV, V, VI |
|  | 4 44 8 | 596 | <.001 | Bilateral paracingulate gyrus  Bilateral anterior cingulate cortex  Bilateral frontal pole  Medial prefrontal cortex |
|  | -40 -18 16 | 437 | <.001 | Left Heschl’s gyrus  Left planum temporale  Left central opercular cortex  Left parietal operculum cortex  Left insular cortex  Left superior temporal gyrus, anterior division |
|  | 40 -30 16 | 53 | <.001 | Right Heschl’s gyrus  Right planum temporale  Right parietal operculum cortex |
|  | -26 -34 68 | 134 | <.001 | Left postcentral gyrus  Left superior parietal lobule |
|  | -32 -28 -14 | 115 | <.001 | Left hippocampus  Left parahippocampal gyrus, posterior division  Left temporal fusiform cortex, posterior division |
|  | 30 -24 -14 | 60 | <.001 | Right hippocampus  Right parahippocampal gyrus, posterior division |
|  | 8 -26 0 | 70 | <.001 | Right thalamus  Brainstem |
|  | -8 -24 -4 | 47 | <.001 | Left thalamus  Brainstem |
|  | -34 -2 60 | 68 | <.001 | Left middle frontal gyrus  Left superior frontal gyrus |
|  | 0 -62 -38 | 51 | <.001 | Vermis VIII/VIIII |
|  | 2 -16 50 | 48 | <.001 | Bilateral supplementary motor cortex  Bilateral precentral gyrus |
|  | -26 -26 70 | 35 | <.001 | Left precentral/postcentral gryi |
|  | 30 -34 68 | 34 | <.001 | Right postcentral gyrus |

*Table S2.* Voxel clusters with significant functional connectivity to right and left precuneus seeds (based on parameters from Harrison et al., 2019) in patients with episodic migraine without accounting for FFMQ total scores

| **Precuneus Seed** | **Cluster Coordinates** | **Cluster Size** | **Cluster Size *p*FDR** | **Cluster Regions** |
| --- | --- | --- | --- | --- |
| Right | 8 -64 18 | 32270 | <.001 | Right precuneus/PCC  Bilateral lateral occipital cortex, superior division  Bilateral lingual gyrus  Bilateral postcentral gyrus  Bilateral intracalcarine cortex  Bilateral occipital pole  Bilateral precentral gyrus  Bilateral cuneal cortex  Bilateral occipital fusiform gyrus  Bilateral anterior cingulate gyrus  Bilateral planum temporale  Bilateral lateral occipital cortex, inferior division  Bilateral superior parietal lobule  Bilateral central opercular cortex  Bilateral cerebellum IV/V/VI  Bilateral insular cortex  Bilateral Heschl’s gyrus  Bilateral temporal occipital fusiform cortex  Right angular gyrus  Vermis IV/V/VI/VII  Bilateral thalamus  Bilateral parietal operculum cortex  Bilateral supracalcarine cortex  Bilateral parahippocampal gyrus, posterior division  Right middle temporal gyrus, temporooccipital  Bilateral supplementary motor cortex  Left temporal fusiform cortex, posterior division  Left putamen  Left hippocampus  Left planum polare  Left superior temporal gyrus, anterior division |
|  | 28 36 34 | 201 | <.001 | Right frontal pole  Right middle frontal gyrus  Right superior frontal gyrus |
| Left | -8 -64 18 | 25078 | <.001 | Left precuneus/PCC  Bilateral lateral occipital cortex, superior division  Bilateral lingual gyrus  Bilateral postcentral gyrus  Bilateral intracalcarine cortex  Bilateral precentral gyrus  Bilateral cuneal cortex  Bilateral occipital pole  Bilateral angular gyrus  Bilateral occipital fusiform gyrus  Left thalamus  Vermis IV/V/VI  Bilateral lateral occipital cortex, inferior division  Bilateral hippocampus  Right planum temporale  Bilateral parahippocampal gyrus, posterior division  Bilateral supracalcarine cortex  Left cerebellum IV/V/VI  Right insular cortex  Bilateral temporal occipital fusiform cortex  Right central opercular cortex  Left temporal fusiform cortex, posterior division  Right middle temporal gyrus, temporooccipital  Right  Right parietal operculum cortex  Right supplementary motor cortex  Right Heschl’s gyrus |
|  | -2 58 -4 | 2718 | <.001 | Left frontal pole  Bilateral paracingulate gyrus  Bilateral anterior cingulate gyrus  Bilateral medial prefrontal cortex |
|  | 26 24 48 | 601 | <.001 | Right middle frontal gyrus  Right superior frontal gyrus  Right frontal pole |
|  | -24 32 44 | 531 | <.001 | Left superior frontal gyrus  Left middle frontal gyrus |
|  | -38 -32 16 | 515 | <.001 | Left Heschl’s gyrus  Left planum temporale  Left parietal operculum cortex  Left insular cortex |
|  | -8 -50 -50 | 247 | <.001 | Bilateral cerebellum VIIII |
|  | -58 -10 -12 | 234 | <.001 | Left middle temporal gyrus, posterior and anterior divisions  Left superior temporal gyrus, posterior division |

*Table S3.* Voxel clusters with significant functional connectivity to NeuroSynth-derived PCC and vmPFC seeds in healthy controls without accounting for FFMQ total scores

| **Seed** | **Cluster Coordinates** | **Cluster Size** | **Cluster Size *p*FDR** | **Cluster Regions** |
| --- | --- | --- | --- | --- |
| PCC | 0 -52 26 | 5143 | <.001 | Bilateral precuneus/PCC  Bilateral hippocampus  Bilateral lingual gyrus  Bilateral anterior cingulate cortex  Bilateral thalamus  Bilateral intracalcarine cortex  Right cuneal cortex  Right parahippocampal gyrus, posterior division  Brainstem  Bilateral cerebellum IV/V  Vermis IV/V |
|  | 4 48 22 | 4090 | <.001 | Bilateral frontal pole  Bilateral paracingulate gyrus  Bilateral superior frontal gyrus  Left middle frontal gyrus  Bilateral medial prefrontal cortex  Bilateral anterior cingulate cortex |
|  | -42 -68 34 | 1338 | <.001 | Left lateral occipital cortex, superior division  Left angular gyrus  Left supramarginal gyrus, posterior division  Left superior parietal lobule |
|  | 48 -62 34 | 1007 | <.001 | Right lateral occipital cortex, superior division  Right angular gyrus |
|  | -62 -42 -4 | 562 | <.001 | Left middle temporal gyrus, anterior, posterior, and temporoccipital divisions  Left superior temporal gyrus, anterior and posterior divisions  Left temporal pole |
|  | 24 36 48 | 426 | <.001 | Right frontal pole  Right superior frontal gyrus  Right middle frontal gyrus |
|  | 62 -12 -8 | 317 | <.001 | Right middle temporal gyrus, anterior, posterior, and temporoccipital divisions  Right superior temporal gyrus, anterior and posterior divisions  Right temporal pole |
|  | 10 -48 -44 | 382 | <.001 | Bilateral cerebellum VIII/IX  Vermis VIII/IX |
|  | -22 -20 -14 | 347 | <.001 | Left hippocampus  Left parahippocampal gyrus, posterior division  Left temporal fusiform cortex, posterior division  Left lingual gyrus |
| vmPFC | 0 50 4 | 8142 | <.001 | Bilateral frontal pole  Bilateral anterior cingulate cortex  Bilateral paracingulate gyrus  Bilateral superior frontal gyrus  Bilateral medial prefrontal cortex  Bilateral middle frontal gyrus  Subcallosal cortex |
|  | -6 -56 22 | 4350 | <.001 | Bilateral precuneus/PCC  Bilateral anterior cingulate cortex  Bilateral lingual gyrus  Bilateral precentral gyrus  Bilateral orbitofrontal cortex  Bilateral caudate  Right thalamus  Bilateral intracalcarine cortex  Bilateral cuneal cortex  Right parahippocampal gyrus, posterior division  Bilateral hippocampus  Brainstem  Bilateral cerebellum IV/V  Vermis IV/V |
|  | -40 -70 32 | 1167 | <.001 | Left lateral occipital cortex, superior division  Left angular gyrus  Left supramarginal gyrus, posterior division |
|  | 48 -64 32 | 1018 | <.001 | Right lateral occipital cortex, superior and inferior divisions  Right angular gyrus  Right middle temporal gyrus, temporooccipital part |
|  | -12 16 0 | 877 | <.001 | Left caudate  Left thalamus  Left orbitofrontal cortex  Left putamen  Left nucleus accumbens  Left insular cortex  Left pallidum |
|  | 24 18 -14 | 813 | <.001 | Right caudate  Right thalamus  Right orbitofrontal cortex  Right putamen  Right nucleus accumbens  Right insular cortex |
|  | 10 -46 -24 | 432 | <.001 | Left hippocampus  Left parahippocampal gyrus, posterior division  Left temporal fusiform cortex, posterior division  Left lingual gyrus  Brainstem  Left vermis IV/V  Bilateral cerebellum III/IV/V/VI |
|  | 14 -98 8 | 429 | <.001 | Bilateral occipital pole  Bilateral lingual gyrus  Right occipital fusiform gyrus  Bilateral intracalcarine cortex  Right cerebellum I/VI  Right lateral occipital cortex, inferior division  Right supracalcarine cortex  Right cuneal cortex |
|  | 54 -32 -8 | 172 | <.001 | Right middle temporal gyrus, posterior and temporoocipital divisions |
|  | -38 -34 10 | 151 | <.001 | Left planum temporale  Left Heschl’s gyrus  Left insular cortex  Left parietal operculum cortex |
|  | -6 -56 -46 | 149 | <.001 | Bilateral cerebellum IX  Right cerebellum VIII  Vermis IX |
|  | -58 -30 -8 | 129 | <.001 | Left middle temporal gyrus, posterior and temporoocipital divisions |
|  | 34 -76 -32 | 129 | <.001 | Right cerebellum crus I/II |
|  | -40 -34 10 | 213 | <.001 | Right planum temporale  Right Heschl’s gyrus  Right insular cortex  Right parietal operculum cortex |
|  | -28 -78 -32 | 114 | <.001 | Left cerebellum crus I/II |

*Table S4.* Voxel clusters with significant functional connectivity to NeuroSynth-derived PCC and vmPFC seeds in patients with episodic migraine without accounting for FFMQ total scores

| **Seed** | **Cluster Coordinates** | **Cluster Size** | **Cluster Size *p*FDR** | **Cluster Regions** |
| --- | --- | --- | --- | --- |
| PCC | 0 -52 26 | 17891 | <.001 | Bilateral precuneus/PCC  Bilateral thalamus  Right cerebellum crus I/II  Brainstem  Bilateral hippocampus  Vermis III/IV/V  Bilateral lingual gyrus  Bilateral caudate  Bilateral parahippocampal gyrus, anterior and posterior divisions  Bilateral cerebellum IV/V  Bilateral anterior cingulate gyrus  Right cerebellum VI/VIIb  Bilateral cuneal cortex  Bilateral temporal fusiform cortex, posterior division  Bilateral amygdala  Bilateral nucleus accumbens  Bilateral intracalcarine cortex  Bilateral supracalcarine cortex  Bilateral occipital pole  Bilateral precentral gyrus  Right temporal pole  Right middle temporal gyrus, anterior, posterior, and temporooccipital divisions  Right orbitofrontal cortex  Right superior temporal gyrus, anterior and posterior divisions  Right inferior temporal gyrus, anterior division  Right temporal and frontal poles  Right occipital fusiform gyrus  Right insular cortex |
|  | 2 46 20 | 11838 | <.001 | Bilateral frontal pole  Bilateral superior frontal gyrus  Bilateral middle frontal gyrus  Bilateral paracingulate gyrus  Bilateral anterior cingulate gyrus  Bilateral medial prefrontal cortex  Left inferior frontal gyrus, pars opercularis  Left inferior frontal gyrus, pars triangularis  Subcallosal cortex |
|  | -60 -40 -4 | 3201 | <.001 | Left middle temporal gyrus, anterior, posterior, and temporoccipital divisions  Left temporal pole  Left orbitofrontal cortex  Left superior temporal gyrus, anterior and posterior divisions  Left inferior temporal gyrus, anterior, posterior, and temporoccipital divisions  Left insular cortex |
|  | -42 -66 30 | 2714 | <.001 | Left lateral occipital cortex, superior division  Left angular gyrus  Left superior parietal lobule  Left supramarginal gyrus, posterior division  Left middle temporal gyrus, temporooccipital part |
|  | 50 -62 28 | 2050 | <.001 | Right lateral occipital cortex, superior and inferior divisions  Right angular gyrus |
|  | -24 -82 -30 | 1012 | <.001 | Left cerebellum crus I/II  Left occipital fusiform gyrus  Left lingual gyrus |
|  | 0 -52 -44 | 846 | <.001 | Bilateral cerebellum VIII/IX  Vermis VIII/IX |
|  | -44 -14 28 | 377 | <.001 | Left precentral and postcentral gyri |
|  | 30 -28 58 | 279 | .002 | Right precentral and postcentral gyri |
| vmPFC | -2 50 4 | 41357 | <.001 | Bilateral precuneus/PCC  Bilateral frontal pole  Bilateral anterior cingulate gyrus  Bilateral middle temporal gyrus, anterior, posterior, and temporoccipital divisions  Bilateral paracingulate gyrus  Bilateral superior frontal gyrus  Bilateral orbitofrontal cortex  Bilateral cerebellum crus I/II  Bilateral occipital pole  Bilateral middle frontal gyrus  Bilateral hippocampus  Bilateral medial prefrontal cortex  Bilateral temporal pole  Bilateral lateral occipital cortex, superior and inferior divisions  Bilateral thalamus  Brainstem  Subcallosal cortex  Bilateral caudate  Bilateral cerebellum IV/V/VI/VIIb/VIII/IX  Bilateral parahippocampal gyrus, anterior and posterior division  Bilateral lingual gyrus  Bilateral putamen  Bilateral hippocampus  Bilateral occipital fusiform gyrus  Bilateral insular cortex  Vermis III/IV/V/VI/VII/VIII/IX  Bilateral inferior temporal gyrus, anterior division  Left inferior temporal gyrus, posterior and temporoocippital divisions  Bilateral nucleus accumbens  Bilateral angular gyrus  Bilateral nucleus accumbens  Bilateral pallidum  Bilateral postcentral gyrus  Bilateral precentral gyrus  Bilateral intracalcarine cortex  Bilateral amygdala  Bilateral cuneal cortex  Left temporal fusiform cortex, posterior and anterior divisions  Bilateral superior temporal gyrus, anterior and posterior divisions  Bilateral inferior frontal gyrus, pars triangularis  Right supracalcarine cortex  Left frontal, central, parietal operculum cortex  Left Heschl's gyrus  Left planum temporale  Left supramarginal gyrus, posterior division  Left temporal fusiform cortex, posterior division |
|  | -48 -62 34 | 2198 | <.001 | Left lateral occipital cortex, superior division  Left angular gyrus  Left supramarginal gyrus, posterior division |
|  | 50 -56 32 | 1662 | <.001 | Right lateral occipital cortex, superior and inferior divisions  Right angular gyrus  Right middle temporal gyrus, temporooccipital part |
|  | -38 -28 16 | 274 | .003 | Left Heschl's gyrus  Left planum temporale  Left central and parietal operculum cortex  Left insular cortex |

*Table S5.* Correlations between FFMQ and PCS subscale scores with clinical pain variables in patients

| Variable | 1 | 2 | 3 | 4 | 5 | 6 | 7 | 8 | 9 | 10 |
| --- | --- | --- | --- | --- | --- | --- | --- | --- | --- | --- |
| 1. PCS-Helplessness |  |  |  |  |  |  |  |  |  |  |
| 2. PCS-Magnification | .52** |  |  |  |  |  |  |  |  |  |
| 3. PCS-Rumination | .67** | .61** |  |  |  |  |  |  |  |  |
| 4. FFMQ-Observing | -0.03 | 0.06 | -0.06 |  |  |  |  |  |  |  |
| 5. FFMQ-Describing | 0.00 | -0.01 | 0.01 | .32** |  |  |  |  |  |  |
| 6. FFMQ-Acting with Awareness | -0.01 | -0.05 | -0.10 | -0.02 | .33** |  |  |  |  |  |
| 7. FFMQ-Non-Judgment | 0.02 | -0.18 | -0.11 | -0.16 | .32** | .46** |  |  |  |  |
| 8. FFMQ-Non-Reactivity | -0.05 | -0.01 | -0.05 | .37** | .31** | 0.07 | .21* |  |  |  |
| 9. Headache Pain Severity | .32** | 0.17 | 0.07 | -0.10 | 0.02 | 0.18 | 0.01 | -0.19 |  |  |
| 10. Headache Impact | .37** | .37** | .37** | -0.04 | -0.02 | -0.15 | -0.14 | -0.19 | .39** |  |
| 11. Headache Frequency | 0.01 | 0.18 | 0.01 | 0.06 | -0.06 | -0.12 | -.29** | 0.09 | -0.08 | 0.00 |

*Note.* * indicates *p* < .05. ** indicates *p* < .01.

*Table S6.* FFMQ subscale scores used to predict NeuroSynth-derived PCC and vmPFC seed-based functional connectivity in patients with episodic migraine

| **Subscale** | **Cluster Coordinates** | **Cluster Size** | **Cluster Size *p*FDR** | **Cluster Regions** |
| --- | --- | --- | --- | --- |
| **vmPFC as a Seed** | | | | |
| Nonjudgment | -26, -76, 48 | 781 | .000 | Left superior lateral occipital cortex  Left angular gyrus |
|  | 4, -58, 44 | 669 | .000 | Precuneus cortex  Posterior cingulate gyrus |
|  | 42, -60, 36 | 581 | .000 | Right superior lateral occipital cortex  Right angular gyrus |
|  | -10, -104, 0 | 421 | .000 | Bilateral occipital pole  Left lingual gyrus  Left intracalcarine cortex |
| **PCC as a Seed** | | | | |
| Nonjudgment | 2, 46, 0 | 128 | .047 | Anterior cingulate gyrus  Bilateral paracingulate gyrus |

*Table S7.* Correlations among FFMQ subscales, PCS subscales, and experimental pain variables in healthy controls

| Variable | 1 | 2 | 3 | 4 | 5 | 6 | 7 | 8 | 9 | 10 |
| --- | --- | --- | --- | --- | --- | --- | --- | --- | --- | --- |
| 1. PCS-Helplessness |  |  |  |  |  |  |  |  |  |  |
| 2. PCS-Magnification | .64** |  |  |  |  |  |  |  |  |  |
| 3. PCS-Rumination | .61** | .51** |  |  |  |  |  |  |  |  |
| 4. FFMQ-Observing | .01 | .21 | 0.14 |  |  |  |  |  |  |  |
| 5. FFMQ-Describing | .18 | .26 | -0.1 | 0.27 |  |  |  |  |  |  |
| 6. FFMQ-Acting with Awareness | -.06 | .03 | 0.02 | 0.21 | 0.18 |  |  |  |  |  |
| 7. FFMQ-Non-Judgment | .15 | -.05 | 0.19 | -0.12 | 0.17 | 0.21 |  |  |  |  |
| 8. FFMQ-Non-Reactivity | .08 | .25 | 0.2 | .69** | .48** | 0.11 | 0.2 |  |  |  |
| 9. Experimental Pain Severity | .09 | -.10 | 0.09 | -0.05 | -0.03 | 0.10 | 0.00 | -0.10 |  |  |
| 10. Experimental Pain Unpleasantness | .11 | -.06 | 0.07 | 0.07 | 0.07 | 0.05 | -0.10 | -0.00 | .93** |  |
| 11. Heat Pain Threshold | -.49** | -.44** | -0.3 | -0.03 | -0.06 | -0.10 | 0.00 | 0.04 | -.55** | -.50** |

*Note.* * indicates *p* < .05. ** indicates *p* < .01.

*Table S8*. FFMQ subscale scores used to predict NeuroSynth-derived PCC and vmPFC seed-based functional connectivity in healthy controls

| **Subscale** | **Cluster Coordinates** | **Cluster Size** | **Cluster Size *p*FDR** | **Cluster Regions** |
| --- | --- | --- | --- | --- |
| **vmPFC as a Seed** | | | | |
| Nonjudgment | -16, -10, -20 | 133 | .03 | Left amygdala  Left hippocampus  Left anterior parahippocampal gyrus |
| **PCC as a Seed** | | | | |
| Describing | 50, -2, 6 | 197 | .005 | Right central opercular cortex  Right precentral gyrus  Right insular cortex  Right postcentral gyrus  Right planum polare |

**STROBE Statement—checklist of items that should be included in reports of observational studies**

|  | **Item No.** | **Recommendation** |  | **Page # where this item**  **is located:** |
| --- | --- | --- | --- | --- |
| **Title and abstract** | 1 | (*a*) Indicate the study’s design with a commonly used term in the title or the abstract |  | 1 |
|  |  | (*b*) Provide in the abstract an informative and balanced summary of what was done and what was found |  | 2 |
| **Introduction** | | | |  |
| Background/rationale | 2 | Explain the scientific background and rationale for the investigation being reported |  | 3-4 |
| Objectives | 3 | State specific objectives, including any prespecified hypotheses |  | 4 |
| **Methods** | | | |  |
| Study design | 4 | Present key elements of study design early in the paper |  | 4-5, 7 |
| Setting | 5 | Describe the setting, locations, and relevant dates, including periods of recruitment, exposure, follow-up, and data collection |  | 4-5 |
| Participants | 6 | (*a*) *Cohort study*—Give the eligibility criteria, and the sources and methods of selection of participants. Describe methods of follow-up  *Case-control study*—Give the eligibility criteria, and the sources and methods of case ascertainment and control selection. Give the rationale for the choice of cases and controls  *Cross-sectional study*—Give the eligibility criteria, and the sources and methods of selection of participants |  | 5 |
|  |  | (*b*) *Cohort study*—For matched studies, give matching criteria and number of exposed and unexposed  *Case-control study*—For matched studies, give matching criteria and the number of controls per case N/A |  | N/A |
| Variables | 7 | Clearly define all outcomes, exposures, predictors, potential confounders, and effect modifiers. Give diagnostic criteria, if applicable |  | 7-8 |
| Data sources/ measurement | 8* | For each variable of interest, give sources of data and details of methods of assessment (measurement). Describe comparability of assessment methods if there is more than one group |  | 7-14 |
| Bias | 9 | Describe any efforts to address potential sources of bias |  |  |
| Study size | 10 | Explain how the study size was arrived at |  | 4-5 (Parent trial referenced) |

Continued on next page

| Quantitative variables | 11 | Explain how quantitative variables were handled in the analyses. If applicable, describe which groupings were chosen and why |  | 9-14 |
| --- | --- | --- | --- | --- |
| Statistical methods | 12 | (*a*) Describe all statistical methods, including those used to control for confounding |  | 9-14 |
|  |  | (*b*) Describe any methods used to examine subgroups and interactions |  | N/A |
|  |  | (*c*) Explain how missing data were addressed |  | 13 |
|  |  | (*d*) *Cohort study*—If applicable, explain how loss to follow-up was addressed  *Case-control study*—If applicable, explain how matching of cases and controls was addressed  *Cross-sectional study*—If applicable, describe analytical methods taking account of sampling strategy |  |  |
|  |  | (*e*) Describe any sensitivity analyses |  | 19 |
| **Results** | | | | |
| Participants | 13* | (a) Report numbers of individuals at each stage of study—eg numbers potentially eligible, examined for eligibility, confirmed eligible, included in the study, completing follow-up, and analysed |  | Parent trial referenced on page 5 |
|  |  | (b) Give reasons for non-participation at each stage |  | Parent trial referenced on page 5 |
|  |  | (c) Consider use of a flow diagram |  | Parent trial referenced on page 5 |
| Descriptive data | 14* | (a) Give characteristics of study participants (eg demographic, clinical, social) and information on exposures and potential confounders |  | 5-6 |
|  |  | (b) Indicate number of participants with missing data for each variable of interest |  | 14 |
|  |  | (c) *Cohort study*—Summarise follow-up time (eg, average and total amount) |  | N/A |
| Outcome data | 15* | *Cohort study*—Report numbers of outcome events or summary measures over time |  | N/A |
|  |  | *Case-control study—*Report numbers in each exposure category, or summary measures of exposure |  | N/A |
|  |  | *Cross-sectional study—*Report numbers of outcome events or summary measures |  | 14-21 |
| Main results | 16 | (*a*) Give unadjusted estimates and, if applicable, confounder-adjusted estimates and their precision (eg, 95% confidence interval). Make clear which confounders were adjusted for and why they were included |  | 14-21 |
|  |  | (*b*) Report category boundaries when continuous variables were categorized |  | N/A |
|  |  | (*c*) If relevant, consider translating estimates of relative risk into absolute risk for a meaningful time period |  | N/A |

Continued on next page

| Other analyses | 17 | Report other analyses done—eg analyses of subgroups and interactions, and sensitivity analyses |  | 19-21 |
| --- | --- | --- | --- | --- |
| **Discussion** | | | | |
| Key results | 18 | Summarise key results with reference to study objectives |  | 21-23 |
| Limitations | 19 | Discuss limitations of the study, taking into account sources of potential bias or imprecision. Discuss both direction and magnitude of any potential bias |  | 23-24 |
| Interpretation | 20 | Give a cautious overall interpretation of results considering objectives, limitations, multiplicity of analyses, results from similar studies, and other relevant evidence |  | 21-25 |
| Generalisability | 21 | Discuss the generalisability (external validity) of the study results |  | 24 |
| **Other information** | |  | | |
| Funding | 22 | Give the source of funding and the role of the funders for the present study and, if applicable, for the original study on which the present article is based |  | 1 |
